# Clinical evaluation of deep learning-based risk profiling in breast cancer histopathology and comparison to an established multigene assay

**DOI:** 10.1101/2023.08.31.23294882

**Authors:** Yinxi Wang, Wenwen Sun, Emelie Karlsson, Sandy Kang Lövgren, Balázs Ács, Mattias Rantalainen, Stephanie Robertson, Johan Hartman

**Affiliations:** Department of Medical Epidemiology and Biostatistics, Karolinska Institutet, Stockholm, Sweden; Stratipath AB, Stockholm, Sweden; Department of Oncology-Pathology, Karolinska Institutet, Stockholm, Sweden; Department of Clinical Pathology and Cancer Diagnostics, Karolinska University Hospital, Stockholm, Sweden; MedTechLabs, BioClinicum, Karolinska University Hospital, Stockholm, Sweden

## Abstract

A significant proportion of oestrogen receptor (ER)-positive and human epidermal growth factor receptor 2 (HER2)-negative early breast cancer patients are categorised as intermediate risk based on classic clinicopathological variables, thus providing limited information to guide treatment decisions. The Prosigna assay is one of the established prognostic multigene assays in clinical practice for risk profiling. Stratipath Breast is a novel deep learning-based image analysis tool that utilises haematoxylin and eosin (HE)-stained histopathological images for risk profiling. In this study, we aimed to evaluate the Stratipath Breast tool for image-based risk profiling and compare it with the Prosigna assay. In a real-world breast cancer case series comprising 234 invasive tumours from patients with early ER+/HER2-breast cancer, clinically intermediate risk and eligible for chemotherapy, clinicopathological data including Prosigna results and corresponding HE-stained tissue slides were retrieved. The digitised HE slides were analysed by Stratipath Breast. Our findings showed that the Stratipath Breast analysis identified 49.6% of the clinically intermediate tumours as low risk and 50.4% as high risk. The Prosigna assay classified 32.5%, 47.0% and 20.5% tumours as low, intermediate and high risk, respectively. Among Prosigna intermediate-risk tumours, 47.3% were stratified as Stratipath low risk and 52.7% as high risk. In addition, 89.7% of Stratipath low-risk cases were classified as Prosigna low/intermediate risk. The overall agreement between the two tests for low-risk and high-risk groups was 71.0%, with a Cohen’s kappa of 0.42. For both risk profiling tests, grade and Ki67 differed significantly between risk groups. In conclusion, for the first time, we here present the results from a clinical evaluation of image-based risk stratification and show a considerable agreement to an established gene expression assay in routine breast pathology. The findings demonstrate that image-based risk profiling may aid in the identification of low-risk patients who could potentially be spared adjuvant chemotherapy.

## INTRODUCTION

Based on classic clinicopathological variables, a significant proportion of oestrogen receptor (ER)-positive and human epidermal growth factor receptor 2 (HER2)-negative early-stage breast cancer are categorised as clinically intermediate risk, thus providing limited information to guide adjuvant chemotherapy decisions. Prognostic risk profiling has become an integrated part of modern breast cancer diagnostics to provide additional risk information for this patient group for identifying patients where adjuvant chemotherapy can be omitted^1-3^.

Among the established prognostic multiparameter diagnostic assays based on gene expression^4^, the Prosigna assay (Prosigna Breast Cancer Prognostic Gene Signature Assay, Veracyte, South San Francisco, USA) is widely used and endorsed by national and international guidelines^5-8^. The Prosigna assay identifies intrinsic molecular subtypes (i.e., luminal A, luminal B, HER2-enriched and basal-like) and provides an individual risk of recurrence (ROR) score between 0 and 100 along with a three-tier risk category (low, intermediate, high) based on ROR score and nodal status. The Prosigna assay contributes with prognostic information for patients with early ER+/HER2-breast cancer and its efficacy has been demonstrated in several study populations^1,9-14^.

The diagnostic foundation with pathological assessment of the well-established prognostic variables such as tumour size, stage and tumour grade, is still an essential part of clinical decision making^15^. Among these, histological grade is one of the most important prognostic factors for breast cancer^16,17^. Approximately 50% of all^17-20^, and around 60% of ER-positive/HER2-negative^1,21^ breast cancers are classified as histological grade 2, which is a heterogenous group of tumours with variations in terms of aggressiveness and prognosis^22,23^, thus, associated with limited value to guide decisions on choice of therapy. A limitation in clinical decision-making is, despite the use of prognostic multigene assays, that tests such as Prosigna may classify up to 44% of histological grade 2 tumours as intermediate risk^24^, which does not add any clinically actionable information. In addition, the diagnostic multigene assays for breast cancer risk stratification show discordances in risk categorisation between different tests^12,25^.

Digital pathology workflows are becoming standard practice and enable application of advanced image analysis in the clinical setting^26^. In addition, the recent evolution in deep learning, a field of artificial intelligence (AI), has further expanded the utility of machine learning techniques in computational pathology, making it possible to predict patient prognosis^27-29^, response to neoadjuvant therapy^30^ or underlying molecular phenotypes^28,31-34^ in breast cancer using computer-based models to analyse and characterise histopathology whole slide images. Hence, computational pathology also plays a central role in precision medicine^26^. By leveraging grade-associated morphological features from haematoxylin and eosin (HE)-stained histopathology slides, deep learning-based image analysis has been shown to enable stratification of grade 2 tumours into two risk groups associated with risk of recurrence^27^.

The novel AI-based precision diagnostic solution, Stratipath Breast (Stratipath AB, Solna, Sweden), is a commercial CE-IVD marked deep learning-based image analysis tool that utilises digitised histopathological whole slide images to stratify intermediate risk patients in terms of risk of recurrence^29^. The test outputs a two-tier risk category. Compared with multigene assays, deep learning-based techniques have the strength of providing fast and cost-efficient solutions.

In this study, we provide the first clinical evaluation of the AI-based Stratipath Breast tool for image-based risk profiling where we compare it with an established multigene assay for risk stratification in a real-world breast cancer case series of clinically intermediate-risk ER+/HER2-tumours.

## MATERIAL AND METHOD

### Patient inclusion and clinical data retrieval

This retrospective real-world case series consisted of 234 invasive breast tumours from patients with early ER-positive HER2-negative breast cancer, clinically assessed as intermediate-risk tumours and eligible for chemotherapy, diagnosed at Karolinska University Hospital and Södersjukhuset, Stockholm, Sweden. All tumours had therefore previously been analysed with the Prosigna assay in clinical routine at point of diagnosis, between the years 2020 and 2022, to evaluate their risk of recurrence according to the Swedish national guidelines^35^. The cohort has partly been expanded from Kjällquist et al.^24^. The Prosigna assay had been performed at the Department of Clinical Pathology and Cancer Diagnostics, Karolinska University Hospital, on sections from formalin-fixed paraffin-embedded (FFPE) breast cancer tissue blocks, according to the manufacturer’s instructions (Veracyte, South San Francisco, CA, USA) on the nCounter system (NanoString Technologies, Seattle, WA, USA) as part of clinical routine. Clinicopathological data including Prosigna results (intrinsic molecular subtype, ROR score (0-100) and risk group) were retrospectively retrieved from electronic records, along with the corresponding archived HE-stained, and parallel sectioned Ki67-stained, FFPE tissue slides.

Clinicopathological tumour data was retrieved from electronic records. Upon diagnosis, the routine biomarkers ER, progesterone receptor (PR), HER2 and Ki67, were assessed according to national guidelines^35^. Monoclonal rabbit anti-ER (clone SP1), anti-PR (clone 1E2), anti-HER-2/*neu* (clone 4B5) and anti-Ki67 (clone 30-9) antibodies were utilised according to the manufacturer’s instructions (BenchMark ULTRA Staining Module, Ventana Medical Systems, Arizona, USA). A positive ER or PR status was defined as >=10% positive tumour nuclei. HER2 status was first determined by IHC, and tumours with 2+ score were subsequently evaluated for gene amplification by HER2 dual-probe *in situ* hybridisation staining with VENTANA HER2 Dual ISH DNA Probe Cocktail assay (Roche Diagnostics, Rotkreutz, Switzerland) together with VENTANA Silver ISH DNP Detection kit and VENTANA Red ISH DIG detection kit according to the manufacturer’s instructions (BenchMark ULTRA IHC/ISH Staining Module, Ventana Medical Systems, Arizona, USA). Ki67 score was reported as a continuous index that describes the percentage of positively stained tumour nuclei within a hotspot containing a minimum of 200 tumour cells or from year 2022 as a global score across the entire tumour due to changes in the national guidelines.

Exclusion criteria are described in the consort diagram in **Supplementary Figure S1**. Only cases with complete results available from both the Prosigna and Stratipath Breast tests were included in the study comparisons.

### Ki67 global scoring

Due to the change in Ki67 scoring recommendations in 2022, all tumours were re-scored for Ki67 by the global scoring method using the open-source image analysis software QuPath^36^. All original Ki67 stained tissue slides were digitised in-house with a Hamamatsu NanoZoomer XR (Hamamatsu Photonics K.K., Shikuoka, Japan) at 40X magnification (0.226 μm/pixel). A protocol for digital Ki67 global scoring using QuPath was followed, described previously^37-39^ and in accordance with recommendations from the International Ki67 in Breast Cancer Working Group (https://www.ki67inbreastcancerwg.org/; accessed on 18 July, 2023). The analysis was run on the entire invasive tumour area of the whole slide image (WSI) and output as a global Ki67 score (%). A few cases without digitised Ki67 slide available (N=22) were manually evaluated by the global scoring method^40^. The cut-offs applied in the national guidelines were used for three-tier groups: Ki67-low (<6%), Ki67-intermediate (6-29%), and Ki67-high (>29%)^6^.

### Stratipath Breast analysis

Stratipath Breast (Stratipath AB, Solna, Sweden) is a commercial CE-IVD marked deep learning-based image analysis tool for risk stratification of breast cancer patients. HE-stained slides of FFPE tissue sections were retrieved and subsequently digitised in-house with a Hamamatsu NanoZoomer XR at 40X magnification (0.226 μm/pixel). Each HE-stained WSI was analysed by Stratipath Breast (version 1.1). The image analysis model encompasses consecutive steps including quality assessment, cancer detection and risk stratification. Twenty-three images did not meet the intrinsic quality control of the Stratipath Breast analysis and were excluded from subsequent analysis (**Supplementary Figure S1**). In addition, for two cases the WSIs were not available for Stratipath Breast analysis. All Stratipath Breast analysis reports underwent pathologist review according to the instructions for use (User manual Stratipath Breast, Stratipath AB) to verify that adequate tumour area was analysed as part of quality control, and cases that did not meet the requirement were excluded (N=14; **Supplementary Figure S1**). For each case Stratipath Breast provides a two-tier risk group; low risk or high risk, together with a continuous risk score (research use only).

### Statistical analysis

Descriptions of agreements between two risk stratification approaches were reported by the actual number and percentage, and Cohen’s kappa was used for two-group comparisons. The differences in distribution of patients belonging to each risk group, with respect to categorical clinical variables, were evaluated by the Fisher’s Exact test when the minimum number of patients in a subgroup was less than 5, or by the chi-square test otherwise. For comparing differences in continuous variables that were not assumed to be normally distributed, the Mann-Whitney U test (comparison across two groups) and Kruskal-Wallis test (comparison across more than two groups) were used. The correlation between continuous scores were calculated with Spearman correlation. All statistical analyses were 2-sided, and a *P* value of less than 0.05 was regarded as significant. The above statistical analyses were performed in IBM SPSS Statistics (version 28.0; IBM, Armonk, New York, USA). Changes in classification between tests were visualised by Sankey diagrams in https://jsfiddle.net.

## RESULTS

### Patient characteristics

A total of 234 early-stage ER-positive/HER2-negative invasive breast tumours were included in the analyses of this study (**Supplementary Figure S1**). The patients’ clinical characteristics and associated Prosigna results are summarised in **Table 1**. Most of the tumours were invasive carcinoma of no special type (NST) or mixed NST (79.5%) and 17.9% were invasive lobular carcinomas (ILC). The median Ki67 score was 24.5% (range 3.85-75.2%) by digital global scoring method. Out of all included tumours, the Prosigna assay classified 76 (32.5%), 110 (47.0%) and 48 (20.5%) tumours as low, intermediate and high risk, respectively. The median ROR score was 47 with a range from 3 to 84. The Prosigna intrinsic molecular subtypes were distributed as follows: 127 (54.3%) luminal A, 107 (45.7%) luminal B, 0 (0%) HER2-enriched and 0 (0%) basal-like.

**Table 1.** Patient characteristics for all included cases and grade 2 cases.

|  | All<br>Frequency (%) | Grade 2<br>Frequency (%) |
| --- | --- | --- |
| N | 234 | 176 |
| Age, mean (Std Dev) | 65.10 (7.88) | 65.11 (8.155) |
| Primary tumour |  |  |
| No | 5 (2.1%) | 4 (2.3%) |
| Yes | 229 (97.9%) | 172 (97.7%) |
| Bilateral breast cancer |  |  |
| No | 225 (96.2%) | 170 (96.6%) |
| Yes | 9 (3.8%) | 6 (3.4%) |
| Histological grade |  |  |
| Grade 1 | 12 (5.1%) | 0 (0%) |
| Grade 2 | 176 (75.2%) | 176 (100%) |
| Grade 3 | 46 (19.7%) | 0 (0%) |
| ER %, median (range) | 99 (20-100) | 99 (20-100) |
| ER status |  |  |
| Positive | 234 (100%) | 176 (100%) |
| PR %, median (range) | 60 (0-100) | 60 (0-100) |
| PR status |  |  |
| Negative | 56 (23.9%) | 48 (27.3%) |
| Positive | 178 (76.1%) | 128 (72.7%) |
| HER2 status |  |  |
| Negative | 234 (100%) | 176 (100%) |
| Ki67 %, median (range)* | 24.49 (3.85-75.21) | 21.19 (4.0-71.72) |
| Ki67 status* |  |  |
| Low | 5 (2.1%) | 3 (1.7%) |
| Intermediate | 151 (64.5%) | 126 (71.6%) |
| High | 78 (33.3%) | 47 (26.7%) |
| Tumour size |  |  |
| <=20mm | 166 (70.9%) | 122 (69.3%) |
| >20mm | 67 (28.6%) | 53 (30.1%) |
| N/A | 1 (0.4%) | 1 (0.6%) |
| pT status |  |  |
| pT1 | 166 (70.9%) | 122 (69.3%) |
| pT2 | 62 (26.5%) | 48 (27.3%) |
| pT3 | 5 (2.1%) | 5 (2.8%) |
| N/A | 1 (0.4%) | 1 (0.6%) |
| Lymph node status |  |  |
| Negative | 211 (90.2%) | 158 (89.8%) |
| Positive | 20 (8.5%) | 16 (9.1%) |
| N/A | 3 (1.3%) | 2 (1.1%) |
| pN status |  |  |
| pN0 | 211 (90.2%) | 158 (89.8%) |
| pN1 | 19 (8.1%) | 15 (8.5%) |
| pN2 | 1 (0.4%) | 1 (0.6%) |
| N/A | 3 (1.3%) | 2 (1.1%) |
| Histological subtype |  |  |
| ILC | 42 (17.9%) | 39 (22.2%) |
| IMC | 3 (1.3%) | 2 (1.1%) |
| Mixed NST | 7 (3.0%) | 5 (2.8%) |
| NST | 179 (76.5%) | 129 (73.3%) |
| Other | 3 (1.3%) | 1 (0.6%) |
| Prosigna ROR score, median (range) | 47 (3-84) | 45 (3-80) |
| Prosigna risk group |  |  |
| Low | 76 (32.5%) | 66 (37.5%) |
| Intermediate | 110 (47.0%) | 83 (47.2%) |
| High | 48 (20.5%) | 27(15.3%) |
| Prosigna intrinsic subtype |  |  |
| Luminal A | 127 (54.3%) | 109 (61.9%) |
| Luminal B | 107 (45.7%) | 67 (38.1%) |

\*Ki67 global scoring method
ER=oestrogen receptor, HER2=human epidermal growth factor receptor 2, ILC=invasive lobular carcinoma, IMC=invasive mucinous carcinoma, N/A=not available, NST=invasive carcinoma of no special type, PR=progesterone receptor, pN=pathological N stage for regional lymph nodes according to TNM 8, pT=pathological T stage for invasive tumour according to TNM 8 (TNM classification of malignant tumours, 2017, 8<sup>th</sup> Ed), ROR=risk of recurrence.

### Comparison between the tests for risk stratification

The Stratipath Breast analysis identified 116 (49.6%) tumours as low risk and 118 (50.4%) as high risk. Among Prosigna intermediate-risk tumours, 52 (47.3%) were stratified as low risk and 58 (52.7%) as high risk by Stratipath Breast (**Figure 1A, Table 2**). In addition, 24 (31.6%) of the 76 Prosigna low-risk cases were upgraded as high-risk by Stratipath Breast, whereas 12 (25.0%) of the 48 Prosigna high-risk cases were downgraded by Stratipath Breast (**Figure 1B, Table 2-3**). The overall agreement between the two tests for low-risk and high-risk groups was 71.0%, with a Cohen’s kappa of 0.42. Prosigna intermediate-risk results were not included in the overall agreement estimate as it is non-informative for treatment decision making. However, when grouping Prosigna low and intermediate risk together, out of the 116 Stratipath low-risk cases 104 (89.7%) were Prosigna low/intermediate risk and 12 (10.3%) were high risk (**Figure 1C, Supplementary Table S1**).

**Table 2.**
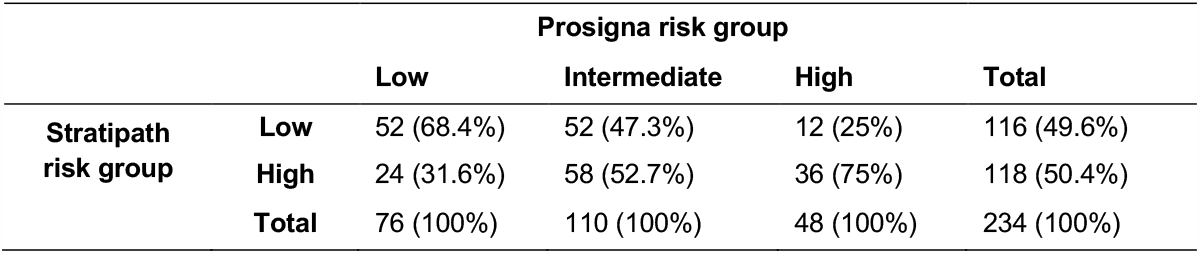
Comparison of agreement in risk stratification between Stratipath Breast risk group and Prosigna risk group.

|  |  | Prosigna risk group |  |  | Total |
| --- | --- | --- | --- | --- | --- |
|  |  | Low | Intermediate | High |  |
| Stratipath risk group | Low | 52 (68.4%) | 52 (47.3%) | 12 (25%) | 116 (49.6%) |
|  | High | 24 (31.6%) | 58 (52.7%) | 36 (75%) | 118 (50.4%) |
|  | Total | 76 (100%) | 110 (100%) | 48 (100%) | 234 (100%) |

**Table 3.** Comparison of agreement in risk stratification between Stratipath Breast risk group and Prosigna risk group for low and high risk only.

|  |  | Prosigna risk group |  |  |
| --- | --- | --- | --- | --- |
|  |  | Low | High | Total |
| Stratipath risk group | Low | 52 (68.4%) | 12 (25%) | 64 (51.6%) |
|  | High | 24 (31.6%) | 36 (75%) | 60 (48.4%) |
|  | Total | 76 (100%) | 48 (100%) | 124 (100%) |

**Figure 1.**
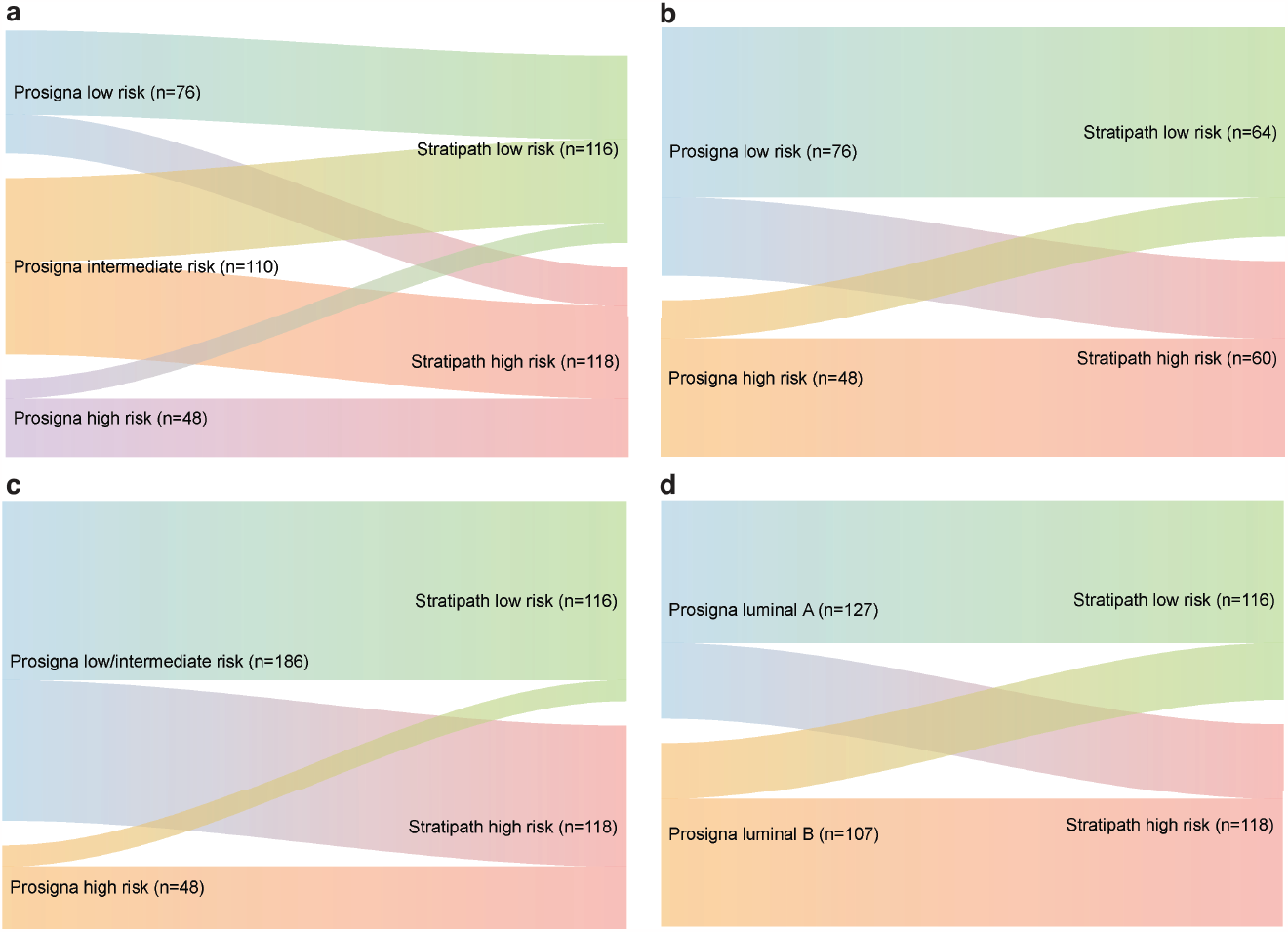
Sankey diagram of the re-classification of risk group between methods. **a** Prosigna risk group vs Stratipath risk group. **b** Prosigna risk group (low and high) vs Stratipath risk group. **c** Prosigna risk group (low/intermediate and high) vs Stratipath risk group. **d** Prosigna intrinsic subtype vs Stratipath risk group.

Among the 176 histological grade 2 tumours, 97 (55.1%) and 79 (44.9%) were stratified as Stratipath low risk and high risk, respectively, whereas 66 (37.5%), 83 (47.2%) and 27 (15.3%) were stratified as Prosigna low, intermediate and high risk, respectively (**Supplementary Table S2**). The agreement between the two tests for low-risk and high-risk groups among grade 2 cases was 68.8%, with a Cohen’s kappa of 0.39. The grade 2 cases showed similar proportions of discordant risk categories as among all cases (**Supplementary Table S3**).

In total 36 cases showed two-level discordant risk category (low and high) by the two tests (**Supplementary Table S4**). There were 24 Stratipath-high, Prosigna-low cases, which all were grade 2 or 3, node negative and luminal A. Among the 12 Stratipath-low, Prosigna-high cases there was a mix of all grades, all but one case was luminal B, and there was a higher proportion of invasive lobular carcinoma (ILC; 33.3%) and invasive mucinous carcinoma (IMC; 16.7%) than in the opposite two-level discordant group (16.7% ILC and 0% IMC). Upon review, two of the Stratipath-low, Prosigna-high cases had incorrectly reported tumour size, which may have altered the ROR score and Prosigna risk category if re-tested.

### Clinicopathological characteristics across Stratipath risk groups

When comparing the distribution of clinical variables in each risk group, there was a significant difference in distribution of grade (p<0.001), Ki67 status (p=0.004), histological subtype (p=0.002) and intrinsic subtype (p<0.001) across Stratipath risk groups, but no difference regarding PR status, lymph node status or tumour size (**Table 4**). The majority of grade 1 (10 of 12) and grade 3 (37 of 46) tumours were stratified as Stratipath low risk and high risk, respectively. There was a significant difference in the distribution of Ki67 score across Stratipath risk groups, with higher Ki67 scores in the high-risk than the low-risk group (Mann-Whitney U test p=0.001; **Figure 2A**). Among grade 2 cases, no difference in Ki67 score was observed between Stratipath risk groups (Mann-Whitney U test p=0.058; **Figure 2B**). For the group of grade 2 tumours, only histological subtype (p=0.010) and intrinsic subtype (p<0.001) differed significantly between the Stratipath risk groups (**Supplementary Table S5**). ILC accounted for 17.9% of the tumours and 29 of the 42 (69.0%) ILC tumours were classified as low risk by Stratipath Breast, with even higher proportion among grade 2 cases (22.2% ILC and 74.4% of ILC as low risk). Among Prosigna intermediate-risk cases, a significant difference between Stratipath low-risk and high-risk groups was identified for grade (p=0.002) and lymph node status (p=0.013; **Supplementary Table S6**).

**Table 4.** Difference of distribution between risk groups for Stratipath Breast and Prosigna per clinicopathological characteristic.

|  | Stratipath risk group |  |  |  | Prosigna risk group |  |  |  |  |
| --- | --- | --- | --- | --- | --- | --- | --- | --- | --- |
|  | Low | High | Total | p-value | Low | Inter-mediate | High | Total | p-value |
| Histological grade |  |  |  | **\*<0.001** |  |  |  |  | **^<0.001** |
| 1 | 10 | 2 | 12 |  | 5 | 5 | 2 | 12 |  |
| 2 | 97 | 79 | 176 |  | 66 | 83 | 27 | 176 |  |
| 3 | 9 | 37 | 46 |  | 5 | 22 | 19 | 46 |  |
| Total | 116 | 118 | 234 |  | 76 | 110 | 48 | 234 |  |
| PR status |  |  |  | **\*0.321** |  |  |  |  | **\*0.011** |
| Negative | 31 | 25 | 56 |  | 26 | 17 | 13 | 56 |  |
| Positive | 85 | 93 | 178 |  | 50 | 93 | 35 | 178 |  |
| Total | 116 | 118 | 234 |  | 76 | 110 | 48 | 234 |  |
| Ki67 status° |  |  |  | **^0.004** |  |  |  |  | **^<0.001** |
| Low | 4 | 1 | 5 |  | 5 | 0 | 0 | 5 |  |
| Intermediate | 84 | 67 | 151 |  | 65 | 70 | 16 | 151 |  |
| High | 28 | 50 | 78 |  | 6 | 40 | 32 | 78 |  |
| Total | 116 | 118 | 234 |  | 76 | 110 | 48 | 234 |  |
| Tumour size |  |  |  | **\*0.918** |  |  |  |  | **\*0.001** |
| <=20mm | 83 | 83 | 166 |  | 45 | 90 | 31 | 166 |  |
| >20mm | 33 | 34 | 67 |  | 31 | 19 | 17 | 67 |  |
| Total | 116 | 117 | 233 |  | 76 | 109 | 48 | 233 |  |
| Lymph node status |  |  |  | **\*0.154** |  |  |  |  | **^0.059** |
| Negative | 102 | 109 | 211 |  | 73 | 98 | 40 | 211 |  |
| Positive | 13 | 7 | 20 |  | 3 | 9 | 8 | 20 |  |
| Total | 115 | 116 | 231 |  | 76 | 107 | 48 | 231 |  |
| Histological subtype |  |  |  | **^0.002** |  |  |  |  | **^0.102** |
| NST | 82 | 97 | 179 |  | 53 | 89 | 37 | 179 |  |
| ILC | 29 | 13 | 42 |  | 21 | 14 | 7 | 42 |  |
| Mixed NST | 2 | 5 | 7 |  | 2 | 4 | 1 | 7 |  |
| IMC | 3 | 0 | 3 |  | 0 | 1 | 2 | 3 |  |
| Other | 0 | 3 | 3 |  | 0 | 2 | 1 | 3 |  |
| Total | 116 | 118 | 234 |  | 76 | 110 | 48 | 234 |  |
| Prosigna subtype |  |  |  | **\*<0.001** |  |  |  |  | **^<0.001** |
| Luminal A | 83 | 44 | 127 |  | 76 | 49 | 2 | 127 |  |
| Luminal B | 33 | 74 | 107 |  | 0 | 61 | 46 | 107 |  |
| Total | 116 | 118 | 234 |  | 76 | 110 | 48 | 234 |  |

\*Chi-Square test; ^Fisher Exact test. All statistical tests are two-sided. °Ki67 global scoring method.
ILC=invasive lobular carcinoma, IMC=invasive mucinous carcinoma, NST=invasive carcinoma of no special type, PR=progesterone receptor.

**Figure 2.**
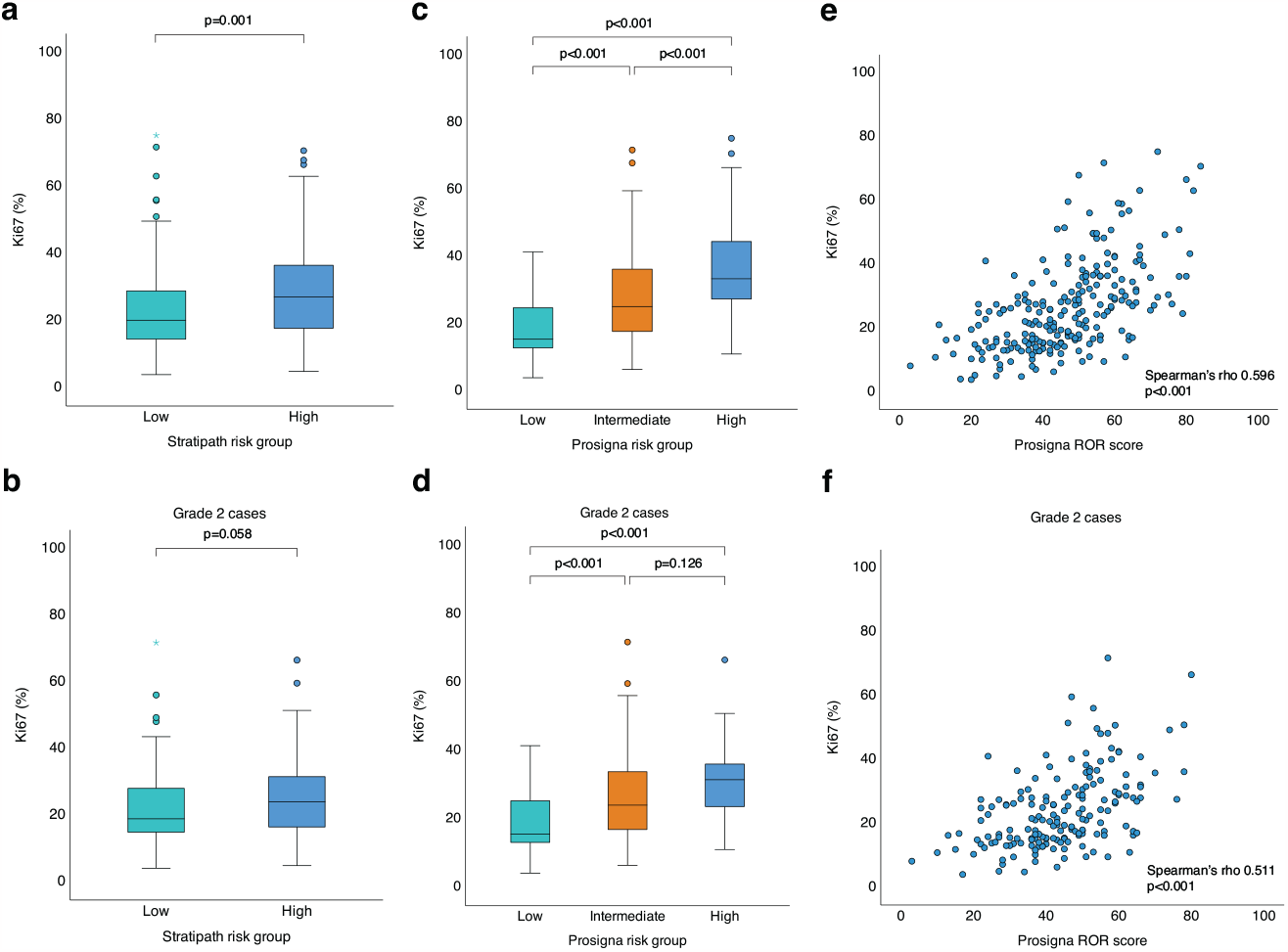
Comparison of Ki67 score across risk groups and correlation to Prosigna risk of recurrence (ROR) score. **a** Significant difference in distribution of Ki67 score across Stratipath Breast risk groups (p=0.001, Mann-Whitney U test). Box plot illustrating median, interquartile range and range. **b** No difference in Ki67 score between Stratipath Breast risk groups among grade 2 cases (p=0.058, Mann-Whitney U test). **c** Significant difference in distribution of Ki67 score across Prosigna risk groups (adjusted p<0.001, between all three groups, Kruskal-Wallis test). **d** Distribution of Ki67 score across Prosigna risk groups for grade 2 cases showed a significant difference between low vs intermediate risk (adjusted p<0.001) and low vs high risk (adjusted p<0.001), but not between intermediate vs high risk (adjusted p=0.126, Kruskal-Wallis test). **e** Significant correlation between Ki67 score and Prosigna ROR score (Spearman’s rho 0.596, p<0.001). **f** Significant correlation between Ki67 score and Prosigna ROR score for grade 2 cases (Spearman’s rho 0.511, p<0.001).

Among the 116 Stratipath low-risk cases, 88 (75.9%) were Ki67-low/intermediate and 28 (24.1%) were Ki67-high (**Supplementary Table S7**). In total, 37.6% of all cases were both Stratipath low risk and Ki67-low/intermediate. Furthermore, 95.5% of the Stratipath low-risk group with Ki67-low/intermediate was Prosigna low or intermediate risk (**Supplementary Table S7**). Turning to the Stratipath high-risk group, two of the 50 (4.0%) Ki67-high cases were classified as Prosigna low risk (**Supplementary Table S8**).

### characteristics across Prosigna risk groups

Across Prosigna risk groups, a significant difference in distribution of grade (p<0.001), PR status (p=0.011), Ki67 status (p<0.001), tumour size (p=0.001) and intrinsic subtype (p<0.001) was observed (**Table 4**) and Ki67 status, tumour size, lymph node status and intrinsic subtype all remained significant among grade 2 cases (**Supplementary Table S5**). There was no difference in the distribution of histological subtype between Prosigna risk groups.

Regarding Ki67 score, a significant difference in distribution of Ki67 score across all Prosigna risk groups was observed (Kruskal-Wallis test adjusted p<0.001; **Figure 2C**). Among grade 2 cases there was a difference in distribution of Ki67 score across the three Prosigna risk groups (Kruskal-Wallis test p<0.001) with a significant difference between low vs intermediate risk (adjusted p<0.001) and low vs high risk (adjusted p<0.001), but not between intermediate vs high risk (adjusted p=0.126; **Figure 2D**). In addition, Ki67 score showed a significant correlation with ROR score (Spearman’s rho 0.596, p<0.001; **Figure 2E**), also among grade 2 cases (Spearman’s rho 0.511, p<0.001; **Figure 2F**).

### ROR score and intrinsic subtype across Stratipath risk groups

ROR scores were higher in the Stratipath high-risk group compared to the low-risk group (p<0.001), across all cases as well as in the Prosigna intermediate-risk group and among grade 2 cases (**Figure 3**). Regarding the distribution of intrinsic subtypes, a total of 83 out of 127 (65.4%) luminal A cases were classified as Stratipath low risk and 74 of 107 (69.2%) luminal B cases as Stratipath high risk (**Figure 1C, Supplementary Table S9**) and similar results were observed among grade 2 cases (**Supplementary Table S10**). A significant difference in distribution of Prosigna intrinsic subtypes across Stratipath risk groups and Prosigna risk groups was identified for all cases as well as for grade 2 cases (Chi-square test and Fisher exact test p<0.001; **Table 4, Supplementary Table S5**).

**Figure 3.**
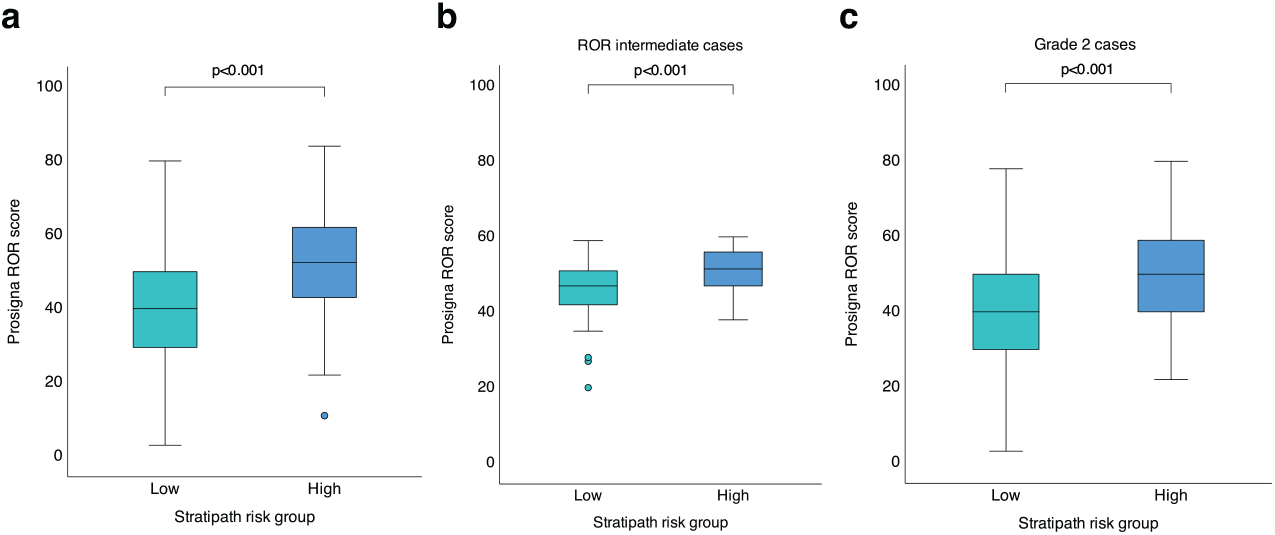
Difference in risk of recurrence (ROR) score across Stratipath risk groups. **a** Higher ROR score in Stratipath high risk group than low risk group (N=234; Mann Whitney U test p<0.001). **b** Higher ROR score in Stratipath high risk group than low risk group among ROR intermediate cases (N=110; Mann Whitney U test p<0.001). **c** Higher ROR score in Stratipath high risk group than low risk group among grade 2 cases (N=176; Mann-Whitney U test p<0.001). Box plots illustrating median, interquartile range and range.

## DISCUSSION

In this study we show the first clinical comparison between the AI-based tool Stratipath Breast and the well-established multigene assay Prosigna for risk profiling of clinically intermediate-risk breast cancer. The agreement between the two tests reached 71.0% (Cohen’s kappa of 0.42) for classifying patients as low risk and high risk. This considerable overall agreement between Stratipath Breast and Prosigna, two methodologically different tests, is on a similar level to what has been observed previously with respect to agreement between different multigene assays^25^. In a comparison of multigene tests in the OPTIMA Prelim trial, the overall agreement between Prosigna risk group and Oncotype DX recurrence score (Exact Sciences, Madison, USA) was 81.0% for low and high risk groups and 77.5% for low/intermediate and high risk groups^25^. A disagreement in this range is expected since these assays are based on different models, gene sets and clinical variables. Although demonstrating robust prognostic value on a population level, discrepancies on the individual patient level are evident when different multigene assays are performed on the same tumour sample^1,12^.

Risk profiling of breast cancer, by e.g., the Prosigna assay, is currently an integrated part of clinical routine diagnostics for clinically intermediate-risk early breast cancer patients. This is crucial to avoid inadequate treatment, especially for ambiguous cases where traditional biomarkers are insufficient to predict if patients would benefit from e.g., additional adjuvant chemotherapy or could be spared chemotherapy. We observed a higher proportion of all cases classified as low risk by Stratipath Breast (49.6%) than by Prosigna (32.5%), which may impact treatment decision to spare patients of chemotherapy. However, as Prosigna also provides an intermediate-risk group, which in this study constituted of almost half of the cases (47%), the risk information remains inconclusive for these patients in guiding treatment decisions. Our findings also showed that Stratipath Breast classified a high proportion (47.3%) of the Prosigna intermediate-risk group as low risk. In addition, the majority (89.6%) of the Stratipath low-risk cases were found in the Prosigna low/intermediate-risk group. The Prosigna high-risk group is generally considered candidates for adjuvant chemotherapy. Furthermore, when assessing Ki67 as a clinical factor together with Stratipath Breast, 95.5% of the Stratipath low-risk with Ki67-low/intermediate cases were also Prosigna low/intermediate risk. This supports the use of additional AI-based risk profiling to identify those patients that can be spared from chemotherapy, assuming that other risk factors are in alignment. Furthermore, differences in intrinsic subtype were observed not only in the Prosigna risk groups but also between Stratipath risk groups with higher proportion of luminal A tumours in the low-risk and luminal B tumours in the high-risk group.

According to Hequet et al, in order to avoid using chemotherapy for one patient, an average of 2.3 Prosigna tests were required, and the cost saving was more significant in lower grade tumours for avoided adjuvant chemotherapy^41^. Special resources at the individual pathology laboratory are required for running the assay on the nCounter platform, including tissue preparation by macrodissection of invasive tumour region and sectioning prior to analyses. In comparison, Stratipath Breast is a fully automated diagnostic tool which operates on routine HE-stained slides, thus, no additional workload except for the digitisation of routine slides is required, ensuring a significantly reduced turnaround time and cost.

The heterogeneous nature of breast cancer and especially inter-tumoural heterogeneity of histological grade 2 tumours has been illustrated by gene expression analysis (DNA microarray), which shows that these tumours constitute of a mixture of gene expression patterns found in grade 1 and 3 tumours^22^. The gene expression signature, Genomic Grade Index, was further developed and has shown prognostic potential to more accurately divide histological grade 2 into a low- and high-risk group associated with risk of recurrence^22,23,42^. Leveraging the capacity of automated feature extraction by deep learning, studies have shown that it is feasible to predict RNA expression profiles^31,43^, DNA mutations^44^, intra-tumoural heterogeneity^45^ or intrinsic subtypes^33^ directly from HE-stained slides. Further, by leveraging grade-related feature extraction, deep learning has shown the ability to stratify grade 2 tumours into a low- and high-risk group^27^.

The AI-based image-analysis tool used in this study extracts information based on the morphological appearance in the HE-stained tumour WSI to determine the patient’s risk category. Histopathological variables including histological subtype and tumour grade showed significant different distribution between low-risk and high-risk groups by Stratipath Breast. The deep learning model has capacity to capture a range of representations/features, that are grade-related, but other than the actual subcomponents routinely identified by the pathologist when determining Nottingham histological grade (i.e., tubular formation, pleomorphism and mitotic count)^27^. Here, we showed that histological grade was associated with both Stratipath Breast risk groups and Prosigna risk groups. The association with tumour grade has been shown for several multigene assays^46-49^ and tumour grade has also been incorporated in prognostic index (Nottingham Px) for prognostic stratification of the clinically intermediate-risk group of breast cancer (node negative ER-positive/HER2-negative)^50^.

To establish the risk category, Stratipath Breast utilises only the WSI as input, i.e., without incorporating other clinical variables. On the contrary, several of the clinical variables shown to be significantly different between Prosigna risk groups in this study, are included in the ROR score. The PAM50 gene expression of the tumour sample designates the intrinsic subtype and is combined with a proliferation score and tumour size to calculate the ROR score^51^. Apart from the apparent methodological difference in the tests, it may be speculated that this along with intra-tumoural heterogeneity could explain the discordances to some extent.

Differences in the prognostic performance between different assays can be explained by several factors, including different molecular markers included in the gene signature assays, where the Prosigna ROR score is largely determined by proliferative features whereas others are driven by ER-related features^49^. We found a significant correlation between the proliferation marker Ki67 and ROR score (Spearman’s rho 0.596) in this clinical case series, which is in line with previous findings^24^. A significant association between Prosigna risk categories and Ki67 status was observed in all patients, and in low-vs intermediate-risk groups and in low-vs high-risk groups in the grade 2 subgroup. For Stratipath Breast, Ki67 was significantly different between low- and high-risk groups when evaluating all patients but not in the subset of grade 2 tumours. This is not unexpected since the PAM50 gene assay incorporates eleven proliferation relative genes, and while Stratipath Breast does not explicitly include any information on proliferation, the AI-based approach has the capacity to capture proliferation associated morphological patterns in the WSIs.

Strengths of this study are that the CE-IVD marked commercial form of both tests was used and in a clinical case series from two sites in the intended patient population. However, the study has several limitations. One limitation to the study is the lack of follow-up information for prognostic comparisons but this was outside of the scope for this study and may instead be evaluated in future studies. Neither was treatment information available for evaluations of the effect on treatment decisions. Another limitation is that the Prosigna assay categorises a relatively large proportion of the cases as intermediate risk, which is non-informative for decision making and was thus excluded in several comparisons focusing on the two-level agreement (low and high risk), and we note that this is an intrinsic limitation of the Prosigna assay.

To conclude, in this study of clinically assessed intermediate-risk ER-positive/HER2-negative breast cancers, we observed a considerable agreement between Prosigna and Stratipath Breast for low-risk and high-risk groups. This is the first study where a commercial multigene assay is compared to the image-based risk profiling tool Stratipath Breast, resulting in a reduced number of intermediate-risk breast cancers. These findings may support in identifying low-risk patients for which chemotherapy could be spared. Further studies with outcome data and impact on treatment decision are of value for clinical comparisons.

## Data Availability Statement

The datasets analysed during the current study are not publicly available due to local privacy laws but are available from the corresponding author upon reasonable request.

## Author contributions

Y.W., M.R., J.H. and S.R. performed study concept and design; Y.W., S.R., M.R. and J.H. performed development of methodology and writing, review, and revision of the paper; S.R. and E.K. provided acquisition and assembly of data; W.S., B.A. and S.K.L. provided analysis; Y.W. and S.R. provided analysis and interpretation of data, and statistical analysis; M.R. and J.H. provided resources and supervised the study. All authors read and approved the final paper.

## Acknowledgements

The work was supported by grants from Swedish Cancer Society, Region Stockholm, MedTechLabs, Cancer Society in Stockholm, Swedish Research Council, VINNOVA, SweLife, Swedish Breast Cancer Association. Stratipath AB provided the Stratipath Breast analysis. B.A. was supported by the Swedish Society for Medical Research postdoctoral grant.

## Ethics Approval and Consent to Participate

The study was approved by the Swedish Ethical Review Authority (2019-01908 with amendment 2021-00017, 2022-01102-02). The study was performed in accordance with the Declaration of Helsinki. No additional informed consent was required in accordance with ethical approval in this non-interventional collection and analysis of data from patient records.

## Supplementary Information

The online version contains supplementary material.

