## Supplementary material for "Clinical evaluation of deep learning-based risk profiling in breast cancer histopathology and comparison to an established multigene assay"

##### Supplementary Figures

**Figure S1.** CONSORT diagram of study cohort.

##### Supplementary Tables

**Table S1.** Comparison of agreement in risk stratification between Stratipath Breast risk group and Prosigna risk group (low/intermediate vs high risk).
**Table S2.** Comparison of agreement in risk stratification between Stratipath Breast risk group and Prosigna risk group for grade 2 cases.
**Table S3.** Comparison of agreement in risk stratification between Stratipath Breast risk group and Prosigna risk group for low and high risk only among grade 2 cases**.**
**Table S4.** Cases with discordant risk category (low and high) between the two risk profiling tests.
**Table S5.** Difference of distribution between risk groups for Stratipath Breast and Prosigna per clinicopathological characteristic for grade 2 cases.
**Table S6.** Difference of distribution among Prosigna intermediate-risk cases between Stratipath Breast risk groups per clinicopathological characteristic.
**Table S7.** Crosstabulation of Ki67 status and Prosigna risk group for Stratipath Breast low-risk cases (N=116).
**Table S8.** Crosstabulation of Ki67 status and Prosigna risk group for Stratipath Breast high-risk cases (N=118).
**Table S9.** Comparison of agreement in risk stratification between Stratipath Breast risk group and Prosigna intrinsic subtype.
**Table S10.** Comparison of agreement in risk stratification between Stratipath Breast risk group and Prosigna intrinsic subtype for grade 2 cases.

#### Supplementary Figures

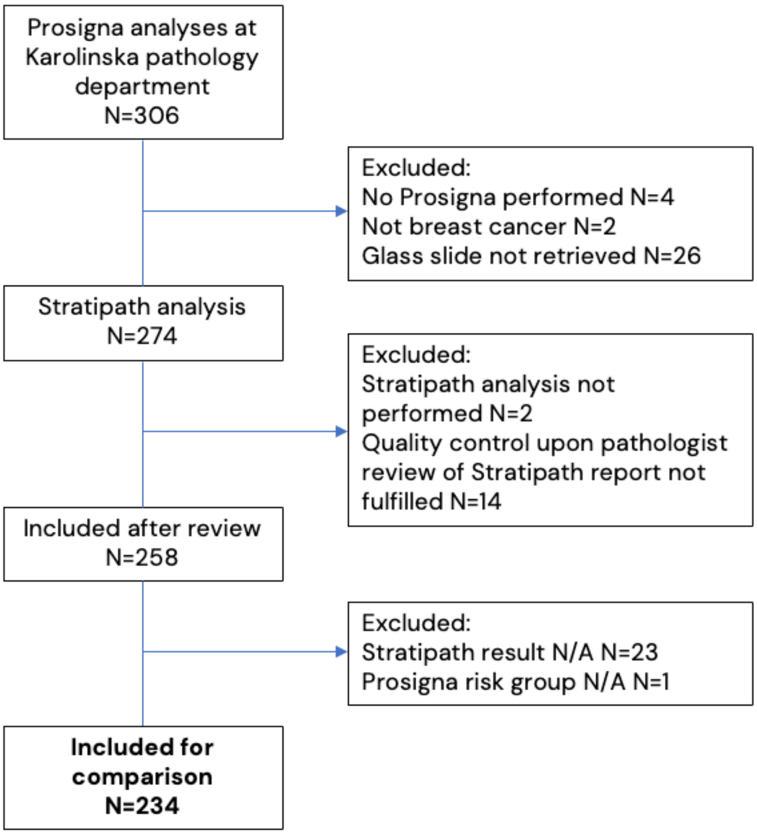


**Figure S1.** CONSORT diagram of study cohort.

### Supplementary Tables

**Table S1.** Comparison of agreement in risk stratification between Stratipath Breast risk group and Prosigna risk group (low/intermediate vs high risk).

| | | Prosigna risk group | | |
|---|---|---|---|---|
| | | **Low/Intermediate** | **High** | **Total** |
| **Stratipath risk group** | **Low** | 104 (55.9%) | 12 (25.0%) | 116 (49.6%) |
| | **High** | 82 (44.1%) | 36 (75.0%) | 118 (50.4%) |
| | **Total** | 186 (100%) | 48 (100%) | 234 (100%) |

**Table S2.** Comparison of agreement in risk stratification between Stratipath Breast risk group and Prosigna risk group for grade 2 cases.

| | | Prosigna Risk group | | | |
|---|---|---|---|---|---|
| | | **Low** | **Intermediate** | **High** | **Total** |
| **Stratipath risk group** | **Low** | 45 (68.2%) | 44 (53.0%) | 8 (29.6%) | 97 (55.1%) |
| | **High** | 21 (31.8%) | 39 (47.0%) | 19 (70.4%) | 79 (44.9%) |
| | **Total** | 66 (100%) | 83 (100%) | 27 (100%) | 176 (100%) |

**Table S3.** Comparison of agreement in risk stratification between Stratipath Breast risk group and Prosigna risk group for low and high risk only among grade 2 cases**.**

| | | Prosigna risk group | | |
|---|---|---|---|---|
| | | **Low** | **High** | **Total** |
| **Stratipath risk group** | **Low** | 45 (68.2%) | 8 (29.6%) | 53 (57.0%) |
| | **High** | 21 (31.8%) | 19 (70.4%) | 40 (43.0%) |
| | **Total** | 66 (100%) | 27 (100%) | 93 (100%) |

**Table S4.** Cases with discordant risk category (low and high) between the two risk profiling tests.

| | Stratipath low \| Prosigna high | Stratipath high \| Prosigna low |
|---|---|---|
| N | 12 | 24 |
| Histological grade | | |
| 1 | 1 (8.3%) | 0 (0%) |
| 2 | 8 (66.7%) | 21 (87.5%) |
| 3 | 3 (25.0%) | 3 (12.5%) |
| Histological subtype | | |
| NST | 6 (50.0%) | 19 (79.2%) |
| ILC | 4 (33.3%) | 4 (16.7%) |
| Mixed NST | 0 (0%) | 1 (4.2%) |
| IMC | 2 (16.7%) | 0 (0%) |
| Other | 0 (0%) | 0 (0%) |
| Lymph node status | | |
| Negative | 10 (83.3%) | 24 (100%) |
| Positive | 2 (16.7%) | 0 (0%) |
| Ki67 status* | | |
| Low | 0 (0%) | 1 (4.2%) |
| Intermediate | 4 (33.3%) | 21 (87.5%) |
| High | 8 (66.7%) | 2 (8.3%) |
| Ki67 % median (range) | 36.2 (17.0-75.2) | 15.6 (4.8-41.4) |
| Prosigna subtype | | |
| Luminal A | 1 (8.3%) | 24 (100%) |
| Luminal B | 11 (91.7%) | 0 (0%) |
| ROR score median (range) | 66.50 (52-80) | 36.50 (11-40) |

*Ki67 global scoring method.
ILC=invasive lobular carcinoma, IMC=invasive mucinous carcinoma, NST=invasive carcinoma of no special type.

**Table S5.** Difference of distribution between risk groups for Stratipath Breast and Prosigna per clinicopathological characteristic for grade 2 cases.

| | **Stratipath risk group** | | | | **Prosigna risk group** | | | | |
|---|---|---|---|---|---|---|---|---|---|
| | **Low** | **High** | **Total** | p-value | **Low** | **Inter-mediate** | **High** | **Total** | p-value |
| PR status | | | | *0.599 | | | | | *0.074 |
| Negative | 28 | 20 | 48 | | 22 | 16 | 10 | 48 | |
| Positive | 69 | 59 | 128 | | 44 | 67 | 17 | 128 | |
| Total | 97 | 79 | 176 | | 66 | 83 | 27 | 176 | |
| Ki67 status° | | | | ^0.230 | | | | | **^<0.001** |
| Low | 2 | 1 | 3 | | 3 | 0 | 0 | 3 | |
| Intermediate | 74 | 52 | 126 | | 57 | 56 | 13 | 126 | |
| High | 21 | 26 | 47 | | 6 | 27 | 14 | 47 | |
| Total | 97 | 79 | 176 | | 66 | 83 | 27 | 176 | |
| Tumour size | | | | *0.264 | | | | | ***0.035** |
| <=20mm | 71 | 51 | 122 | | 40 | 65 | 17 | 122 | |
| >20mm | 26 | 27 | 53 | | 26 | 17 | 10 | 53 | |
| Total | 97 | 78 | 175 | | 66 | 82 | 27 | 175 | |
| Lymph node status | | | | *0.928 | | | | | **^<0.001** |
| Negative | 87 | 71 | 158 | | 64 | 75 | 19 | 158 | |
| Positive | 9 | 7 | 16 | | 2 | 6 | 8 | 16 | |
| Total | 96 | 78 | 174 | | 66 | 81 | 27 | 174 | |
| Histological subtype | | | | **^0.010** | | | | | ^0.271 |
| NST | 64 | 65 | 129 | | 45 | 65 | 19 | 129 | |
| ILC | 29 | 10 | 39 | | 20 | 13 | 6 | 39 | |
| Mixed NST | 2 | 3 | 5 | | 1 | 3 | 1 | 5 | |
| IMC | 2 | 0 | 2 | | 0 | 1 | 1 | 2 | |
| Other | 0 | 1 | 1 | | 0 | 1 | 0 | 1 | |
| Total | 97 | 79 | 176 | | 66 | 83 | 27 | 176 | |
| Prosigna subtype | | | | ***<0.001** | | | | | **^<0.001** |
| Luminal A | 71 | 38 | 109 | | 66 | 41 | 2 | 109 | |
| Luminal B | 26 | 41 | 67 | | 0 | 42 | 25 | 67 | |
| Total | 97 | 79 | 176 | | 66 | 83 | 27 | 176 | |

*Chi-Square test; ^Fisher Exact test. All statistical tests are two-sided. °Ki67 global scoring method.
ILC=invasive lobular carcinoma, IMC=invasive mucinous carcinoma, NST=invasive carcinoma of no special type, PR=progesterone receptor.

**Table S6.** Difference of distribution among Prosigna intermediate-risk cases between Stratipath Breast risk groups per clinicopathological characteristic.

| | **Stratipath risk group** | | | |
|---|---|---|---|---|
| | **Low** | **High** | **Total** | p-value |
| Histological grade | | | | **^0.002** |
| 1 | 4 | 1 | 5 | |
| 2 | 44 | 39 | 83 | |
| 3 | 4 | 18 | 22 | |
| Total | 52 | 58 | 110 | |
| PR status | | | | *0.985 |
| Neg | 8 | 9 | 17 | |
| Pos | 44 | 49 | 93 | |
| Total | 52 | 58 | 110 | |
| Ki67 status | | | | *0.248 |
| Intermediate | 36 | 34 | 70 | |
| High | 16 | 24 | 40 | |
| Total | 52 | 58 | 110 | |
| Tumour size | | | | *0.297 |
| <=20mm | 45 | 45 | 90 | |
| >20mm | 7 | 12 | 19 | |
| Total | 52 | 57 | 109 | |
| Lymph node status | | | | **^0.013** |
| Neg | 43 | 55 | 98 | |
| Pos | 8 | 1 | 9 | |
| Total | 51 | 56 | 107 | |
| Histological subtype | | | | ^0.432 |
| NST | 42 | 47 | 89 | |
| ILC | 8 | 6 | 14 | |
| Mixed NST | 1 | 3 | 4 | |
| IMC | 1 | 0 | 1 | |
| Other | 0 | 2 | 2 | |
| Total | 52 | 58 | 110 | |

*Chi-Square test; ^Fisher Exact test. All statistical tests are two-sided. ILC=invasive lobular carcinoma, IMC=invasive mucinous carcinoma, NST=invasive carcinoma of no special type, PR=progesterone receptor.

**Table S7.** Crosstabulation of Ki67 status and Prosigna risk group for Stratipath Breast low-risk cases (N=116).

| | | Ki67 status | | |
|---|---|---|---|---|
| | | **Low/Intermediate (<=29%)** | **High (>29%)** | **Total** |
| **Prosigna risk group** | **Low** | 48 (54.55%) | 4 (14.3%) | 52 (44.83%) |
| | **Intermediate** | 36 (40.90%) | 16 (57.1%) | 52 (44.83%) |
| | **High** | 4 (4.55%) | 8 (28.6%) | 12 (10.34%) |
| | **Total** | 88 (100%) | 28 (100%) | 116 (100%) |

**Table S8.** Crosstabulation of Ki67 status and Prosigna risk group for Stratipath Breast high-risk cases (N=118).

| | | Ki67 status | | |
|---|---|---|---|---|
| | | **Low/Intermediate (<=29%)** | **High (>29%)** | **Total** |
| **Prosigna risk group** | **Low** | 22 (32.4%) | 2 (4.0%) | 24 (20.3%) |
| | **Intermediate** | 34 (50.0% | 24 (48.0%) | 58 (49.2%) |
| | **High** | 12 (17.6%) | 24 (48.0%) | 36 (30.5%) |
| | **Total** | 68 (100%) | 50 (100%) | 118 (100%) |

**Table S9.** Comparison of agreement in risk stratification between Stratipath Breast risk group and Prosigna intrinsic subtype.

| | | Prosigna subtype | | |
|---|---|---|---|---|
| | | **Luminal A** | **Luminal B** | **Total** |
| **Stratipath risk group** | **Low** | 83 (65.4%) | 33 (30.8%) | 116 (49.6%) |
| | **High** | 44 (34.6%) | 74 (69.2%) | 118 (50.4%) |
| | **Total** | 127 (100%) | 107 (100%) | 234 (100%) |

**Table S10.** Comparison of agreement in risk stratification between Stratipath Breast risk group and Prosigna intrinsic subtype for grade 2 cases.

| | | Prosigna subtype | | |
|---|---|---|---|---|
| | | **Luminal A** | **Luminal B** | **Total** |
| **Stratipath risk group** | **Low** | 71 (65.1%) | 26 (38.8%) | 97 (55.1%) |
| | **High** | 38 (34.9%) | 41 (61.2%) | 79 (44.9%) |
| | **Total** | 109 (100%) | 67 (100%) | 176 (100%) |
